# Cerebral hypoperfusion is a late pathological event in the course of Alzheimer’s disease

**DOI:** 10.1101/2021.07.02.21259911

**Authors:** Khazar Ahmadi, Joana B. Pereira, David Berron, Jacob Vogel, Silvia Ingala, Olof T. Strandberg, Shorena Janelidze, Frederik Barkhof, Josef Pfeuffer, Linda Knutsson, Danielle van Westen, Henk J.M.M. Mutsaerts, Sebastian Palmqvist, Oskar Hansson

**Author notes:** **For correspondence:** (O.H); (K.A). Clinical Memory Research Unit, Department of Clinical Sciences, Lund University, Sölvegatan 19, 22362, Lund, Sweden (Khazar Ahmadi).

## Abstract

Although several studies have shown decreased cerebral blood flow (CBF) in Alzheimer’s disease (AD), the role of hypoperfusion in the disease pathogenesis remains unclear. Combining arterial spin labeling MRI, positron emission tomography, and biomarkers of cerebrospinal fluid, we investigated the associations between CBF and the key mechanisms in AD including amyloid-β (Aβ) and tau pathology, synaptic dysfunction and axonal degeneration. Further, we applied a disease progression modeling to characterize the temporal sequence of different AD biomarkers. Lower perfusion was observed in the temporo-occipito-parietal regions in the Aβ-positive cognitively impaired compared to both the Aβ-positive and Aβ-negative cognitively unimpaired individuals. In participants along the AD spectrum (those with Aβ pathology regardless of their cognitive status), CBF was inversely associated with tau and synaptic dysfunction, but not Aβ, in similar cortical regions. Moreover, the disease progression modeling revealed that CBF disruption followed the abnormality of biomarkers of Aβ, tau and brain atrophy. These findings indicate that tau tangles and synaptic degeneration are more closely connected with CBF changes rather than Aβ pathology. This supports the notion that hypoperfusion is not an early event associated with the build-up of Aβ during the preclinical phase of AD.

## Introduction

Alzheimer’s disease (AD) is a neurodegenerative disorder characterized by the accumulation of amyloid-β (Aβ) plaques and neurofibrillary tangles of hyperphosphorylated tau. These pathological changes precede the onset of dementia by many years, leading to the conceptualization of AD as a continuum evolving from preclinical to prodromal stages and finally to dementia (***Aisen et al., 2017; McKhann et al., 2011***). Multiple neuroimaging studies have shown alterations in functional connectivity (***Berron et al., 2020; Franzmeier et al., 2019; Hahn et al., 2019***), brain metabolism (***Mosconi et al., 2008; Riederer et al., 2018; Verclytte et al., 2016***) and perfusion (***Huang et al., 2018; Mattsson et al., 2014***) in different cortical regions through the course of AD. The latter can be noninvasively measured with arterial spin labeling (ASL) which utilizes magnetically labeled water in the blood as an endogenous tracer (***Williams et al., 1992***). Due to the close coupling of glucose metabolism and cerebral blood flow (CBF), the patterns of regional hypometabolism on well-established positron emission tomography with [18F]-fluorodeoxyglucose (FDG-PET) overlap with reduced CBF on ASL scans (***Riederer et al., 2018; Musiek et al., 2012***). This renders ASL as a promising radiation-free candidate for the diagnosis and monitoring of AD.

Despite several reports on CBF changes in AD (***Binnewijzend et al., 2016; Wolk and Detre, 2012; Fazlollahi et al., 2020***), their association with the core pathologies including Aβ and tau burden remains elusive. In a previous study, a negative correlation was reported between amyloid-PET and ASL-CBF in temporo-parietal regions across the AD continuum (***Mattsson et al., 2014***). In contrast, other studies have suggested a link between cerebrospinal fluid (CSF) biomarkers of tau, but not Aβ, with perfusion abnormalities on single photon emission computed tomography (***Habert et al., 2010; Stomrud et al., 2012***). The few available studies on the relationship between ASL-CBF and tau-PET have yielded mixed findings. While one study has found negative associations in widespread temporo-parietal regions (***Albrecht et al., 2020***), another one has shown that higher tau-PET uptake is correlated with hypoperfusion exclusively in the entorhinal cortex (***Rubinski et al., 2021***). Moreover, no studies to date have investigated the associations of ASL-CBF with synaptic and axonal markers in the CSF. There is also the important question whether decreased CBF triggers the chain of pathological events in AD, or if it is a consequence of the downstream effects of Aβ and tau. Thus, the overarching aims of the current study were to examine the multimodal relationships between ASL-CBF with distinct imaging and CSF biomarkers across the AD spectrum and to identify when along the disease evolution ASL-CBF changes occur.

To this end, we first measured ASL-CBF variations in Aβ-positive or negative cognitively unimpaired individuals and cognitively impaired patients with Aβ pathology. We then evaluated the association between CBF and deposition of Aβ and tau measured with Aβ-PET and CSF Aβ42/40, tau-PET and CSF phosopho-tau217 (P-tau217), as well as the CSF concentrations of synaptic (Neuronal Pentraxin2 to total tau ratio; NPTX2/T-tau), and axonal (neurofilament light chain; NfL) biomarkers. Finally, we ran a disease progression modeling (***Young et al., 2018***) approach to estimate the order of abnormality of these markers. Specifically, we addressed the following questions. i) When does CBF change in the AD continuum? ii) Which brain regions are primarily affected in AD by CBF alterations? iii) Are CBF changes associated with Aβ or tau burden in the AD spectrum? iv) Are CBF variations corelated with markers of synaptic or axonal integrity? Our results demonstrate that CBF alterations occur at later stages of the AD continuum. These changes are most pronounced in the occipito-temporo-parietal areas and are largely associated with tau load and synaptic degeneration.

## Results

### Participants and methodology overview

The present study comprised 137 cognitively unimpaired (CU) controls, and 119 cognitively impaired (CI) patients diagnosed either with mild cognitive impairment or AD with dementia. The groups were further stratified into Aβ-positive and negative individuals based on the CSF Aβ42/40 ratio. Demographic and clinical characteristics of the participants are summarized in Table 1.

**Table 1.** Demographic summary. Data are presented as mean values followed by (standard deviations). Demographic factors were compared between groups using *X*^2^ and ANOVA tests. MMSE = Mini-Mental State Examination. ADAS-cog = Alzheimer’s Disease Assessment Scale-Cognitive subscale. Out of 119 CI patients, 55 had mild cognitive impairment and 64 had AD with dementia.

| | Aβ-negative CU<br>N = 94 | Aβ-positive CU<br>N = 43 | Aβ-positive CI<br>N = 119 | P value |
| --- | --- | --- | --- | --- |
| Males/females | 45/49 | 23/20 | 49/70 | 0.331 |
| Age | 60.07 (7.80) | 68.81 (7.70) | 72.90 (6.86) | < 0.0001 |
| Years of education | 13.47 (3.78) | 12.83 (3.96) | 12.71 (4.44) | 0.398 |
| APOE ε4 carriers | 30.9% | 74.4% | 73.9% | < 0.0001 |
| MMSE | 29.04 (1.12) | 28.81 (1.40) | 23.66 (4.39) | < 0.0001 |
| ADAS-Cog delayed word recall errors | 2.09 (1.52) | 2.98 (2.06) | 8.03 (2.05) | < 0.0001 |

Voxel-wise and region of interest (ROI)-based general linear models were performed to assess between-group differences in CBF and its relationship with the PET and CSF measures. The ROI-based analyses were conducted in six a priori defined ROIs including lateral and medial parts of the parietal and temporal lobes, middle frontal gyrus and superior lateral occipital cortex (Figure 1). All the analyses were adjusted for age, sex and ASL sequence version (see Methods; MRI acquisition and processing). To cover brain regions affected by tau and Aβ pathology, composite-ROIs were created. Tau-PET ROIs (illustrated in Supplementary Figure 1) approximated Braak staging scheme (***Braak and Braak, 1991***) and encompassed an early (entorhinal) ROI corresponding to Braak stage I-II, a temporal meta-ROI reflecting Braak stage I-IV, and a late neocortical meta-ROI corresponding to Braak stage V-VI (***Ossenkoppele et al., 2018***). The Aβ-PET neocortical composite ROI covered prefrontal, parietal, temporal lateral, anterior/posterior cingulate, and precuneus (***Lundqvist et al., 2013; Landau et al., 2014***).

**Figure 1.**
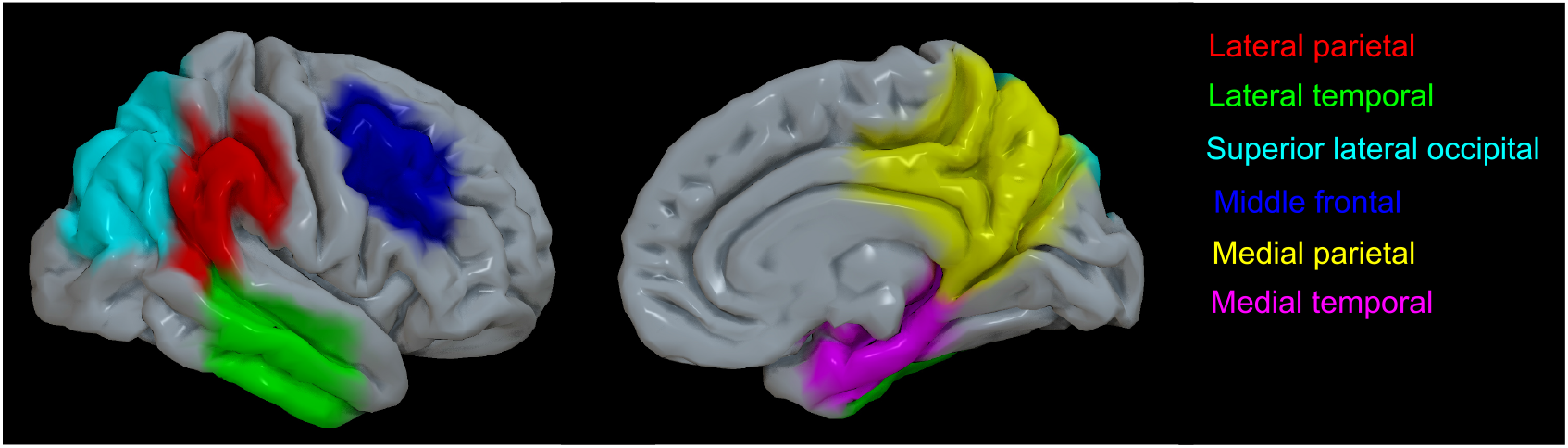
A priori-defined CBF-ROIs projected on the cortical surface. The left and right panels represent the lateral and medial views, respectively. Each ROI is displayed with a distinct color.

To estimate the timing of CBF disruption along the disease pathogenesis, we applied Subtype and Stage Inference (SuStaIn) approach, which combines disease progression modeling with traditional clustering to achieve probabilistic spatiotemporal classification (***Young et al., 2018***). CSF levels of Aβ42/40 and P-tau217, tau-PET deposition in the aforementioned three ROIs, a composite volumetric measure in AD signature regions (***Jack Jr et al., 2017; Ossenkoppele et al., 2018***) and weighted mean CBF in Aβ-negative CU individuals and those on the AD continuum i.e., all Aβ-positive individuals were used as inputs for this modeling framework (see Methods; disease progression modeling).

### CBF differences between diagnostic groups

#### Aβ-negative CU vs. Aβ-positive CU

No significant difference in CBF was observed when comparing Aβ-negative and Aβ-positive CU individuals, neither with the ROI-based nor with the voxel-wise analyses.

#### Aβ-negative CU vs. Aβ-positive CI

The ROI-based analyses revealed significant CBF reductions in the lateral parietal (β = -0.059, *P*_*FDR*_ = 0.0052), superior lateral occipital (β = -0.056, *P*_*FDR*_ = 0.0443) and middle frontal (β = -0.074, *P*_*FDR*_ = 0.0021) ROIs in Aβ-positive CI individuals compared to Aβ-negative CU participants. These findings were further confirmed by the voxel-wise analyses (Figure 2 A), demonstrating decreased CBF in the bilateral superior lateral occipital, bilateral lateral/medial parietal and the left lateral temporal regions (p < 0.05, FWE-corrected).

**Figure 2.**
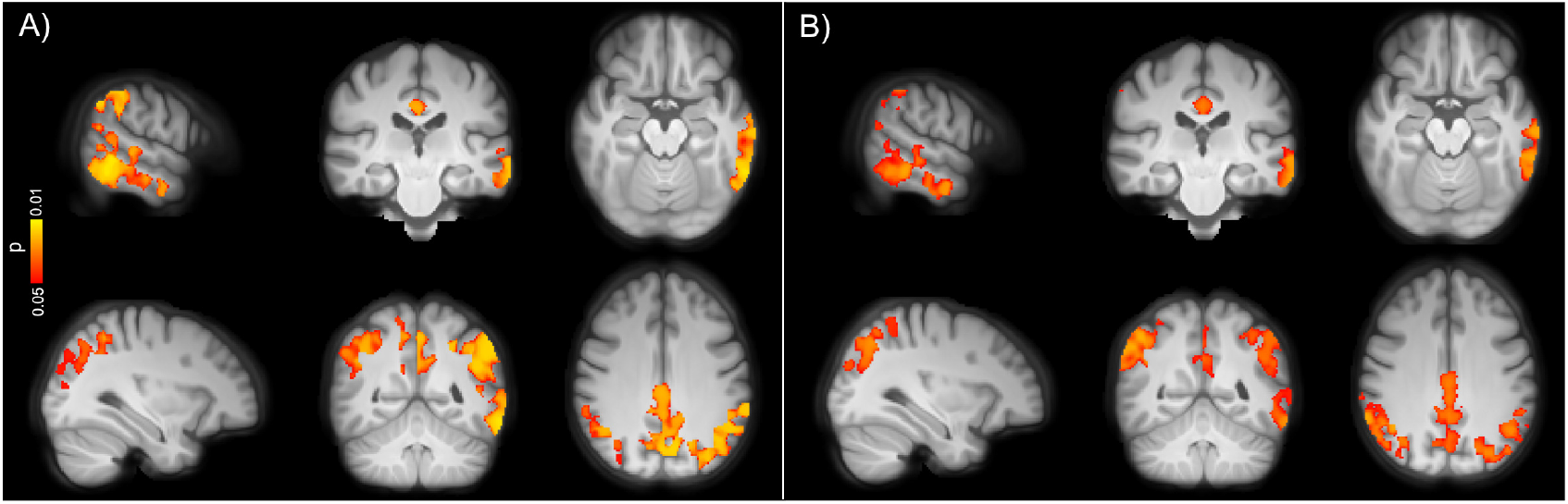
Voxel-wise group differences between A) Aβ-negative CU and B) Aβ-positive CU vs. Aβ-positive CI individuals adjusted for age, sex and ASL sequence version. Clusters with significantly decreased CBF in Aβ-positive CI patients are overlaid on the mean population T1-weighted volume in radiological space (p < 0.05, FWE corrected). The top and bottom panels display different representative slices.

#### Aβ-positive CU vs. Aβ-positive CI

The overall hypoperfusion pattern in Aβ-positive CI with respect to Aβ-positive CU individuals was similar to the pattern observed in the same group when compared to Aβ-negative CU subjects, albeit less pronounced. Specifically, lower CBF was observed in the lateral parietal (β = -0.047, PFDR = 0.0426), superior lateral occipital (β = -0.070, PFDR = 0.0426) and middle frontal (β = -0.050, PFDR = 0.0426) ROIs in Aβ-positive CI individuals. As depicted in Figure 2 B, the voxel-wise analyses showed decreased CBF in the lateral temporal, medial/lateral parietal and superior lateral occipital cortices, largely in line with the ROI-based analyses (p < 0.05, FWE-corrected).

Additional correction for the grey matter (GM) volume did not alter the observed findings between the two CU groups. However, the patterns of decreased CBF in Aβ-positive CI patients compared to both the Aβ-negative and Aβ-positive CU participants were diminished after controlling for the GM volume (see Supplementary Table 1 and Supplementary Figure 2).

### Correlations between CBF and Aβ pathology

Next, we studied the associations between CBF and Aβ pathology measured by Aβ-PET SUVR in the neocortical composite ROI and CSF levels of Aβ42/40. In CU individuals, no associations were observed between CBF and markers of Aβ, neither with the ROI-based nor with the voxel-wise analyses. Similarly, CBF was not correlated with Aβ pathology in individuals along the AD continuum.

### Correlations between CBF and tau pathology

To study associations between CBF and tau pathology, we initially used the tau-PET data. No significant associations were found between CBF and tau load in any of the early, temporal and late tau-PET ROIs in CU individuals regardless of Aβ status. Additionally, CBF was not associated with CSF levels of P-tau217 in this group. In individuals on the AD spectrum, however, both the ROI-based and voxel-wise analyses demonstrated inverse associations between CBF and tau biomarkers. Higher tau-PET uptake in the early ROI was significantly associated with decreased CBF in the lateral parietal (β = -0.052, *P*_*FDR*_ = 0.0134), lateral temporal (β = -0.054, *P*_*FDR*_ = 0.0134), and superior lateral occipital (β = -0.081, *P*_*FDR*_ = 0.0068) ROIs. Tau burden in the temporal meta-ROI was negatively associated with CBF in the lateral parietal (β = -0.060, *P*_*FDR*_ = 0.0001), lateral temporal (β = -0.064, *P*_*FDR*_ = 0.0001), the superior lateral occipital (β = -0.088, *P*_*FDR*_ = 0.00005) and middle frontal (β = -0.034, *P*_*FDR*_ = 0.0499) ROIs. Moreover, tau load in the late meta-ROI had inverse associations with CBF in the same regions [lateral parietal (β = -0.122, *P*_*FDR*_ = 0.00005), lateral temporal (β = -0.086, *P*_*FDR*_ = 0.0046), superior lateral occipital (β = -0.128, *P*_*FDR*_ = 0.0011) and middle frontal (β = -0.087, *P*_*FDR*_ = 0.0046) ROIs]. The associations of CBF with tau-PET ROIs were still significant after controlling for CSF concentrations of Aβ42/40 (Supplementary Table 2). Analogous findings were observed after controlling for cognitive status and GM volume (see Supplementary Table- and Figure 3). The overall significant associations between CBF and tau uptake in each of the three ROIs are plotted in Supplementary Figure 4. Further using the voxel-wise analyses, similar patterns of hypoperfusion were observed for all three tau-PET ROIs mainly in the occipito-temporo-parietal regions (Figure 3). Except for the tau-PET SUVRs in the early ROI, additional adjustment for the cognitive status did not alter the observed voxel-wise associations, as shown in Supplementary Figure 5. This also held true when controlling for GM volume (Supplementary Figure 6).

**Figure 3.**
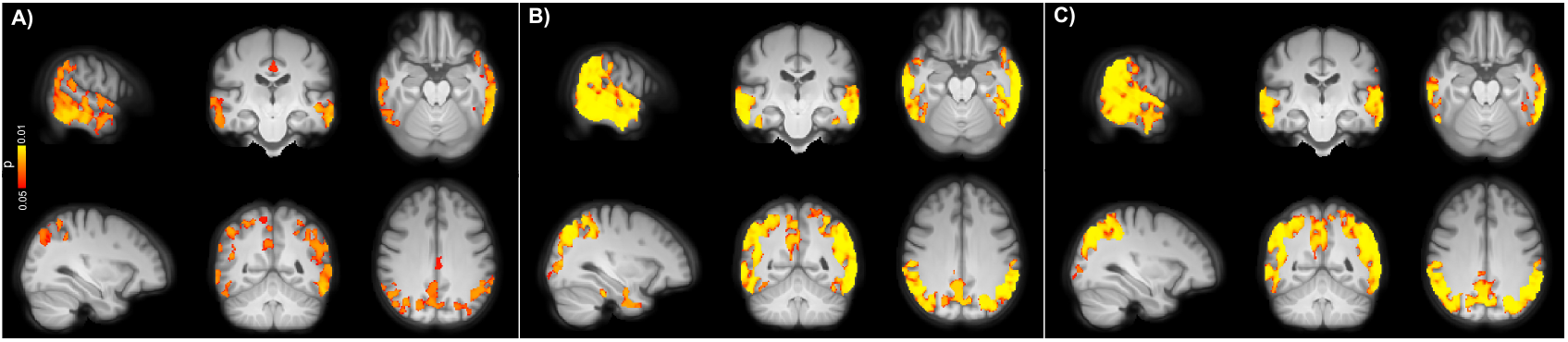
Voxel-wise associations between CBF and tau uptake in A) early, B) temporal and C) late ROIs in the AD spectrum adjusted for age, sex and ASL sequence version. CBF reduction was primarily observed in lateral temporal, lateral/medial parietal and superior lateral occipital regions (p < 0.05, FWE corrected). Conventions as for Figure 2.

Afterwards, we examined the associations between CBF and CSF P-tau217 which is an earlier marker than tau PET (***Mattsson-Carlgren et al., 2020***). Elevated CSF levels of P-tau217 were correlated with lower CBF in lateral parietal (β = -9.643e-05, *P*_*FDR*_ = 0.0117), lateral temporal (β = -8.518e-05, *P*_*FDR*_ = 0.0381), superior lateral occipital (β = -1.708e-04, *P*_*FDR*_ = 0.0017) and middle frontal (β = -8.738e-05, *P*_*FDR*_ = 0.0381), see Supplementary Figure 7. After adjustment for GM volume, only the associations in the lateral parietal and superior lateral occipital remained significant [β = -8.18e-05, p = 0.0185, and β = -1.65e-04, *P*_*FDR*_ = 0.0049, respectively]. Similarly, the voxel-wise analyses showed a negative correlation between higher levels of P-tau217 and hypoperfusion in the occipito-temporo-parietal areas (Figure 4). As depicted in Supplementary Figure 8, following further adjustment for the GM volume, the observed voxel-wise associations were restricted to clusters within the lateral temporal and superior lateral occipital cortex.

**Figure 4.**
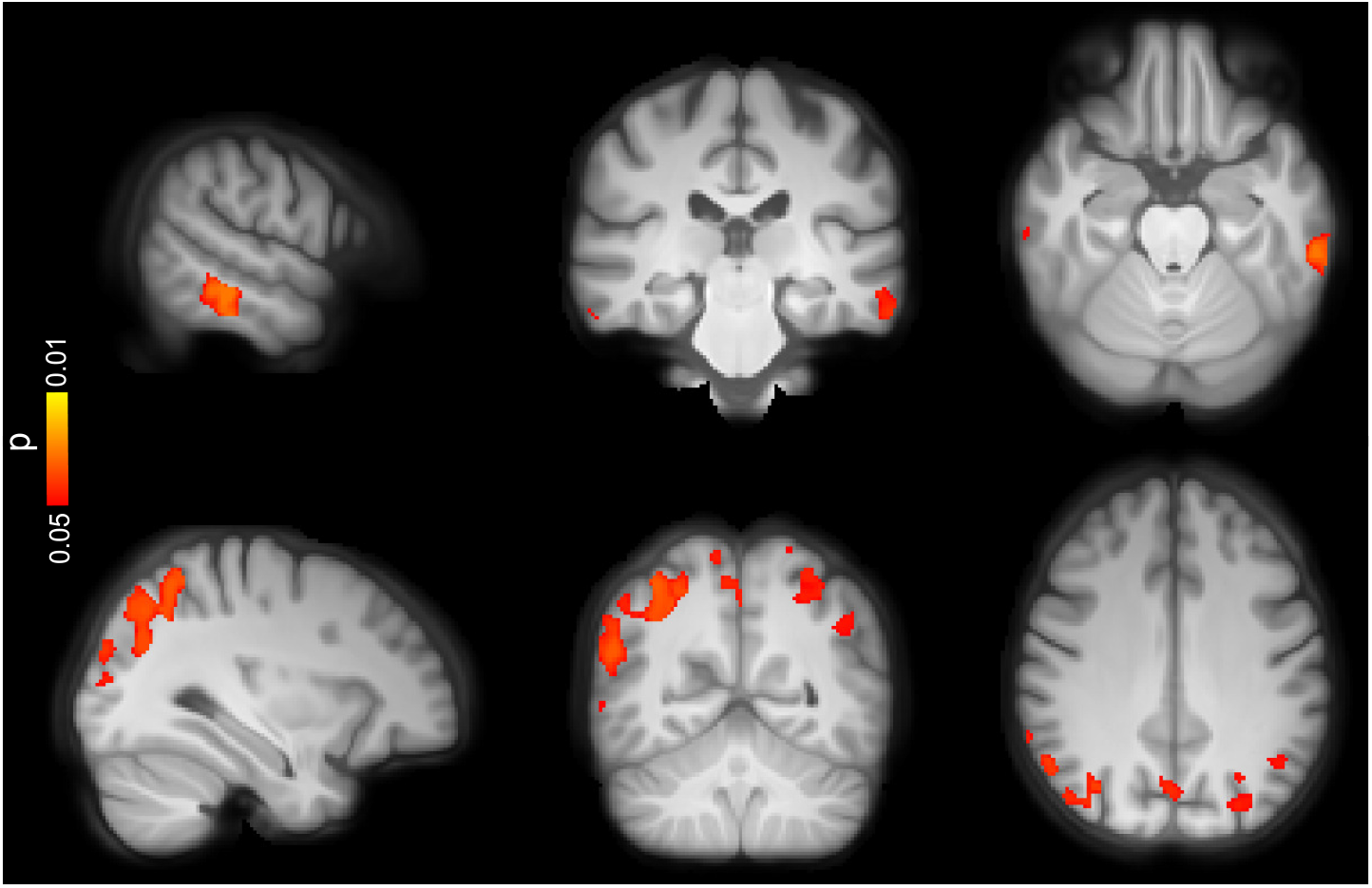
Voxel-wise associations between CBF and CSF concentrations of P-tau217 in the AD spectrum adjusted for age, sex and ASL sequence version. Patterns of hypoperfusion were observed in the same trajectory as for tau-PET albeit to a lesser extent (p < 0.05, FWE corrected). Conventions as for Figure 2.

### Correlations between CBF and CSF biomarkers of synaptic and axonal integrity

To study the associations between CBF and markers of neurodegeneration, we used CSF levels of NfL, which increases in relation to ongoing axonal degeneration (***Gaetani et al., 2019***), and NPTX2/T-tau ratio which decreases with respect to synaptic degeneration (***Galasko et al., 2019***). None of the ROI-based and voxel-wise analyses revealed any significant correlation between CBF and CSF levels of NfL or NPTX2/T-tau in CU individuals. In contrast, a positive association was observed between NPTX2/T-tau and CBF in the AD spectrum for lateral parietal (β = 0.062, PFDR = 0.0069), lateral temporal (β = 0.064, PFDR = 0.0072), superior lateral occipital (β = 0.096, PFDR = 0.0028) and middle frontal (β = 0.074, PFDR = 0.0028) ROIs, see Supplementary Figure 9. The observed correlations were largely corroborated by the voxel-wise analyses showing hypoperfusion with decreasing concentrations of NPTX2/T-tau (see Figure 5). Additional correction for the GM volume did not affect the observed ROI-based and voxel-wise associations (Supplementary Figure 10). Conversely, no significant associations were found between NfL and CBF in this group, neither with the ROI-based nor with the voxel-wise analyses.

**Figure 5.**
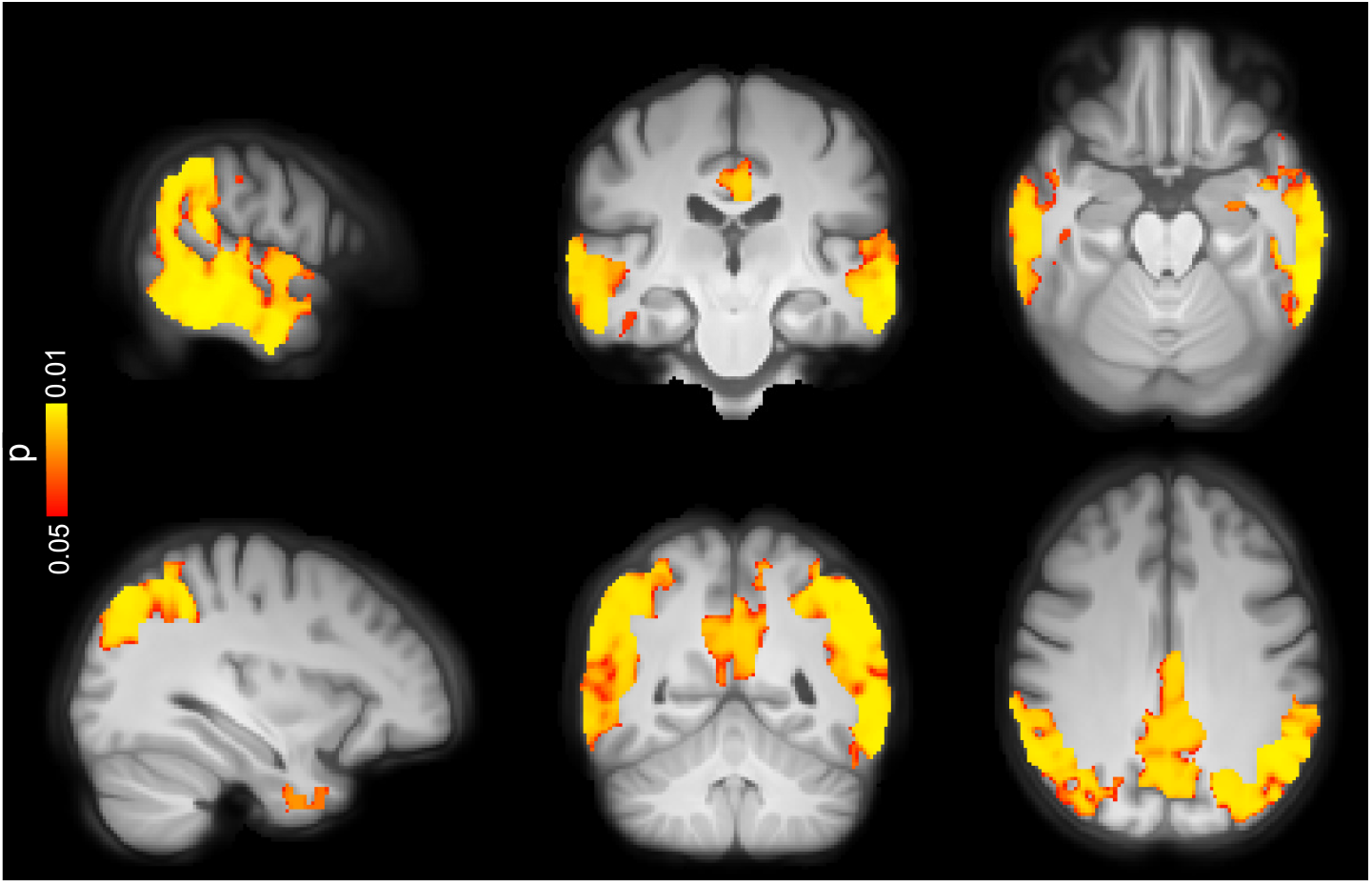
Voxel-wise associations between CBF and CSF levels of NPTX2/T-tau in the AD spectrum adjusted for age, sex and ASL sequence version. Decreasing levels of NPTX/T-tau were accompanied by CBF reduction mainly in occipito-temporo-parietal regions (p < 0.05, FWE corrected). Conventions as for Figure 2.

### Staging of CBF with respect to biomarkers of Aβ, tau and neurodegeneration

SuStaIn was applied to probabilistically assign individuals in Aβ-negative CU group and those along the AD continuum to one of 21 progressive stages (see Figure 6). In accordance with the aforementioned results, the model predicted that abnormal CSF concentrations of Aβ42/40 and P-tau217 initiated the progression of the disease. This was followed by pathological deposition of tau-PET in early and temporal/limbic ROIs, as well as neurodegeneration indicated by the atrophy in the AD signature regions and abnormal tau burden in the late meta-ROI. Only subsequent to these changes, did ASL-CBF alterations become apparent.

**Figure 6.**
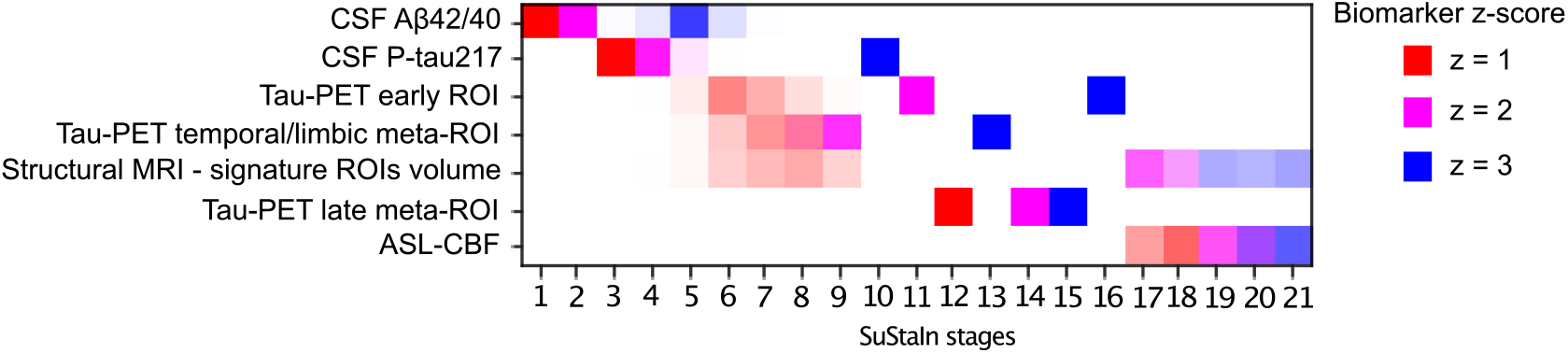
Progression pattern of AD predicted by SuStaIn. While the earliest disease stages comprised abnormalities in CSF markers of Aβ and tau, CBF variations occurred during the latest stages of the disease progression. At each stage, the accumulative probability for each biomarker going from one z-score to another is indexed by distinct colors i.e., red, magenta and blue representing z-scores of 1, 2 and 3, respectively. Higher opacity indicates more confidence in the ordering. Note that in both CU individuals and those along the AD continuum, the weighted mean CBF was obtained in occipito-temporo-parietal regions demonstrating significant voxel-wise associations with tau-PET burden in the early ROI in the AD spectrum (see Figure 3 A).

## Discussion

The present study disentangles the multimodal associations of ASL-CBF with different pathological markers in AD. We found that i) CBF did not differ between the Aβ-negative and Aβ-positive CU groups. However in Aβ-positive CI patients, CBF was markedly reduced compared to CU individuals independent of Aβ pathology; ii) the CBF reduction were mainly localized in the occipito-temporoparietal regions; iii) CBF was not associated with markers of Aβ neither in CU nor in individuals on the AD continuum; iv) higher tau-PET uptake and increased concentrations of CSF P-tau217 were correlated with hypoperfusion patterns in the AD spectrum; v) abnormal levels of the NPTX2/T-tau ratio, a marker of synaptic degeneration, were associated with lower CBF in the same group; and vi) pathological reduction of CBF was evident only after the abnormality of Aβ, tau and regional cortical atrophy. Taken together, these findings indicate that ASL-CBF changes are not an early event associated with aggregation of Aβ plaques during the preclinical phase of AD. Instead, these alterations are more tightly linked to tau pathology, and synaptic degeneration.

The observed CBF differences between Aβ-positive CI and the CU groups are consistent with the accumulating evidence demonstrating widespread hypoperfusion across different cortical regions in AD (***Dai et al., 2009; Johnson et al., 2005; Binnewijzend et al., 2013***). When additional correction for GM volume was applied, the observed effect size was decreased suggesting that between group CBF differences may not be fully independent of GM atrophy. Conflicting results have been reported on CBF changes in CU individuals with elevated risk of AD. While some studies have shown patterns of hyperperfusion in CU APOE ε4 carriers (***Wierenga et al., 2014; McKiernan et al., 2020***), others have found no CBF changes in CU individuals with Aβ pathology (***Binnewijzend et al., 2016***). Our findings endorse the latter indicating that ASL-CBF is a marker of the disease severity, not directly related to Aβ deposition.

Lack of an association between CBF and Aβ burden, as suggested by our results, is in line with a previous report on non-demented humans with unilateral occlusion of precerebral arteries, showing that long-lasting cerebral hypoperfusion does not result in aggregation of Aβ (***Hansson et al., 2018***). Another study, however, has found positive associations between hyperperfusion and increased Aβ-PET load in a small sample (N = 13) of Aβ-positive CU individuals (***Fazlollahi et al., 2020***). On the other hand, inverse associations between lower perfusion and increased Aβ-PET uptake have been reported across the spectrum of sporadic AD and in asymptomatic and mildly symptomatic subjects with autosomal dominant AD (***Mattsson et al., 2014; McDade et al., 2014***). While the observed discrepancy suggests that the potential role of tau should be taken into account when assessing the CBF-Aβ relationship, it may also arise from the differences in the sample size or acquisition of the ASL-CBF images. In the current study, we applied the recommended implementation for the clinical use of ASL i.e., pseudo-continuous sequence with background suppression and 3D gradient- and spin-echo readout, whereas in the latter two studies, a pulsed sequence was used, which is less sensitive and more prone to being influenced by the variations in arterial transit time (***Alsop et al., 2015***). Our findings on the negative CBF-tau associations mainly in the occipito-temporo-parietal cortex in the AD spectrum are in general agreement with previous studies comparing tau-PET uptake with glucose metabolism using FDG-PET (***Whitwell et al., 2018***), and neuropathological results showing higher tau pathology with increased concentrations of vascular endothelial growth factor, an indicator of hypoxia-induced hypoperfusion (***Thomas et al., 2015***). A recent study with direct comparison of ASL-CBF and tau-PET has also shown CBF reduction in similar cortical regions (***Albrecht et al., 2020***). The underlying mechanisms linking tau pathology to CBF decline in AD are yet to be determined. Evidence from animal models has suggested that chronic lower perfusion promotes the accumulation of tau tangles (***Park et al., 2019; Shimada et al., 2019***). In contrast, it has been proposed that neurofibrillary tau tangles have detrimental effects on the integrity of the cortical microvasculature (***Bennett et al., 2018***).

In this study, the unique combination of imaging and CSF measures allowed us to explore the associations of ASL-CBF not only with Aβ and tau as the hallmarks of AD, but also with synaptic and axonal function. Importantly, we found that lower CBF was associated with lower levels of the NPTX2/T-tau ratio, indicating that synaptic degeneration is accompanied by CBF reduction. NPTX2 is a marker of glutamatergic synaptic plasticity (***Swanson et al., 2016***), which has been shown to be down-regulated in AD correlating with cognitive deterioration and hippocampal volume (***Xiao et al., 2017***). Here, we used NPTX2/T-tau ratio as it has been shown to have a high classification accuracy in discriminating AD from normal controls and a strong correlation with cognition (***Galasko et al., 2019***). Together, these findings suggest that CBF alterations in the AD spectrum are elicited by the spread of tau tangles and synaptic damage.

Contrary to what we expected, no correlation was observed between NfL and ASL-CBF. Although there are no published reports on such associations, a few studies have found a link between higher CSF or plasma levels of NfL with cortical hypometabolism in AD (***Mayeli et al., 2019; Benedet et al., 2019***). Further, another study has reported longitudinal associations between higher concentrations of NfL and decline in several neuroimaging measures including FDG-PET (***Mielke et al., 2019***). However, no cross-sectional correlations were found between either CSF or plasma levels of NfL with any of the neuroimaging measures in that study. This agrees with our observation on the absences of a cross-sectional relationship between ASL-CBF and NfL. Collectively, our findings suggest that variations in CBF are more related to synaptic integrity rather than the axonal damage.

Whether perfusion disruption is a cause or consequence of AD remains unclear (***Austin et al., 2011; Mazza et al., 2011***). Some studies have reported that CBF deficiency induces AD neuropathology (***Iturria-Medina et al., 2016; Zhao and Gong, 2015; Koike et al., 2010***) whereas others have shown that cerebral hypoperfusion is secondary to neurodegeneration (***Hansson et al., 2018***). By modeling the disease progression using SuStaIn, we found that anomalous concentrations of Aβ and tau along with cortical volume loss preceded the CBF disturbances in AD, lending support to the latter view. Nevertheless, it is still of paramount importance to treat cardiovascular risk factors in the elderly population to decrease the risk of vascular dementia and the additive effects of lacunae and white matter lesions as co-pathologies in AD.

The cross-sectional nature of this study did not allow us to determine the temporal relationship of ASL-CBF with each of the imaging and CSF measures. Future studies are, thus, warranted to elucidate the longitudinal associations between CBF and markers of Aβ, tau, synaptic and axonal integrity. Many ASL-studies have used in-house scripts for image processing, which hampers the reproducibility of the results. Here, we employed open source tools aimed at standardization of ASL image processing. However, it should be stressed that due to the limited spatial resolution of ASL, the observed findings may not represent the dynamics of blood flow within the cerebral capillaries.

In conclusion, these results provide in vivo evidence indicating that tau aggregates and neurodegeneration, but not Aβ, are closely connected to ASL-CBF alterations across the AD continuum. This suggests that cerebral hypoperfusion, at least measured by ASL, does not stimulate the development of AD pathology but rather is a late reactive change due to the reduced metabolic demand.

## Methods and Materials

### Participants

The participants were recruited from the Swedish BioFINDER-2 study (NCT03174938), which has been previously described in detail (***Palmqvist et al., 2020; Spotorno et al., 2020***). Out of 447 eligible participants, we included 256 individuals, who were at least 50 years of age and whose ASL scans had passed the quality control procedure (see MRI acquisition and processing). All participants gave written informed consent, and the study was approved by the Ethical Review Board in Lund, Sweden.

### MRI acquisition and processing

In accordance with the recommendations of the white paper (***Alsop et al., 2015***), ASL scans were acquired using a prototype 3D pseudo-continuous (pCASL) sequence with background suppression and gradient- and spin-echo (GRASE) readout in a 3T MAGNETOM Prisma scanner with a 64-channel receiver-coil array (Siemens Healthcare, Erlangen, Germany). The labeling plane was planned using an MR-angiogram and the following parameters were used for the pCASL sequence: TR | TE = 4600 | 21.76 ms, flip angle (FA) = 28°, labeling duration | post-labeling delay = 1500 | 2000 ms, reconstructed voxel size = 1.9 × 1.9 × 4 mm^3^, FoV = 240 mm, GRAPPA factor = 2, Turbo factor = 12, EPI factor = 31, 3 segments, 36 axial slices, 12 pairs of control/label images and total acquisition time of 5:50 min. A proton density weighted (M0) volume with TR = 4000 ms was also acquired to calibrate estimated CBF into absolute units. Halfway through the study, the ASL prototype sequence was updated. The protocol was adjusted to optimally resemble the pre-update version. Additionally, a whole-brain T1-weighted scan (MPRAGE sequence, TR | TE | TI = 1900 | 2.54 | 900 ms, resolution = 1 × 1 × 1 mm^3^, FA = 9°, FoV = 256 × 256 mm^2^, 176 slices, bandwidth = 220 Hz/px TA = 5:15 min) and a T2-weighted FLAIR scan (TR | TE | TI = 5000 | 393 | 1800 ms, band-width = 781 Hz/px TA = 4:37 min, same resolution and FoV as for the T1-weighted image) were obtained. ASL and structural scans were processed using the publicly available ExploreASL tool-box (https://github.com/ExploreASL/ExploreASL) developed in MATLAB which uses the Statistical Parametric Mapping (SPM 12) routines. The processing pipeline has been described in detail elsewhere (***Mutsaerts et al., 2020***). Briefly, white matter hyperintensities (WMH) were segmented on the FLAIR image and used to fill the corresponding hypointensities on T1-weighted scan. Voxel intensities in these lesion regions were replaced by bias-field corrected values from the surrounding normally appearing WM on T1 image. Subsequently, the anatomical image was segmented into grey matter (GM), WM and CSF using computational anatomy toolbox 12 (CAT12) to obtain partial volume (PV) maps which were normalized to the MNI152 space. ASL scans were motion-corrected and co-registered to the T1 scan. The M0 image was smoothed with a 16 mm FWHM Gaussian kernel. CBF was quantified using a single compartment model (***Alsop et al., 2015***) as follows:

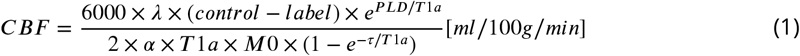

where *λ* is the brain/blood partition coefficient (0.9 ml/g), PLD is post-labeling delay (2000 ms), T1a is the longitudinal relaxation time of arterial blood, *α* is the labeling efficiency (85%), M0 is the proton density image intensity and *τ* is the labeling duration (1500 ms). Subsequently, PV effects were corrected and the CBF images were transformed into MNI152 space. Finally, ASL scans were quality controlled by an automatic method which ranks the processed images based on their spatial coefficient of variation, reflecting the amount of arterial transit time artefacts (***Mutsaerts et al., 2019***). This procedure was reviewed by a trained radiologist (S.I., with 5 years of experience) to detect vascular or other sources of artefacts. Region of interest (ROI) masks were obtained by intersecting the Harvard-Oxford (HO) atlas with individual GM and WM masks. Mean bilateral PV-corrected CBF values were calculated for the atlas ROIs in the standard space (***Mutsaerts et al., 2020***).

### Regions of interest definition for CBF

In line with previous studies, regional CBF values for the atlas ROIs were normalized to CBF of the precentral gyrus to adjust for individual variations in the blood flow (***Mattsson et al., 2014; Yew et al., 2017***). No between group CBF differences were found in this region, prior to the normalization. A set of a priori ROIs were defined based on previous studies showing CBF alterations within these regions in the AD spectrum. These a priori defined ROIs encompassed lateral and medial parts of the parietal and temporal cortex, middle frontal gyrus and superior division of the lateral occipital cortex (Figure 1). The lateral and medial parietal meta-ROIs consisted of anterior/posterior supramarginal gyri, angular gyrus and posterior cingulate plus precuneus, respectively. The lateral temporal meta-ROI included anterior/posterior divisions of the middle and inferior temporal gyri while hippocampus and anterior/posterior divisions of parahippocampal gyrus (anterior parahippocampal gyrus in HO atlas represents the entorhinal cortex) were grouped into the medial temporal meta-ROI. The weighted-mean CBF for a priori ROIs was obtained by the sum of products of the mean CBF in each sub-ROI and its volume, divided by the sum of the volumes of all the sub-ROIs constituting each priori ROI.

### PET acquisition and processing

Tau-PET scans using [18F] RO948 were performed on a GE Discovery MI scanner (General Electric Medical Systems) 70 - 90 minutes post-injection. [18F] flutemetamol Aβ-PET scans were acquired on the same scanner 90 - 110 minutes following the injection as previously described (***Leuzy et al., 2020***). Standardized uptake value ratio (SUVR) images were calculated using the inferior cerebellar GM and pons as the reference regions for tau- and Aβ-PET, respectively. The aforementioned T1-weighted scan was used for PET image co-registration and template normalization. Note that the Aβ-PET data were only available for a subset of 183 participants, including 87 Aβ-negative, 40 Aβ-positive CU individuals and 56 Aβ-positive CI patients. Tau- and Aβ-PET composite ROIs were created using the Freesurfer-based anatomical parcellation (https://surfer.nmr.mgh.harvard.edu/).

### CSF biomarkers

Lumbar CSF sampling and analysis were conducted according to the Alzheimer’s Association Flow Chart (***Blennow et al., 2010***). As previously described, the analysis of P-tau217 assays was performed using the Meso Scale Discovery (MSD) platform (***Janelidze et al., 2020***). Total tau (T-tau) was quantified using Innotest® immunoassay (Fujirebio; Gent, Belgium). CSF Aβ42 and Aβ40 levels were measured using MSD or Fujirebio Lumipulse assays according to the manufacturer’s instructions, and the ratio values were subsequently calculated. NfL was measured using a Simoa kit (Quanterix; Billerica, MA). NPTX2 was measured in the facilities of ADx (Gent, Belgium) using a research grade sandwich ELISA. The number of available data per group for each of the CSF and PET biomarkers, together with the corresponding mean values are shown in Supplementary Table 4.

### Statistical analyses

The ROI-based analyses were performed using R (version 3.5.2). Group differences across demographics were compared using analysis of variance (ANOVA) or chi-squared *X*^2^ tests (table 1). Global CBF differences between the diagnostic groups were evaluated by univariate linear regression models with CBF of a priori ROIs as the dependent variable and the groups as the independent variable. To assess the relationships between CBF and each of the PET and CSF markers of Aβ, tau, synaptic dysfunction and axonal integrity, the regression analysis was separately applied to the AD continuum, and CU participants with the CBF in a priori ROIs as the dependent variable and each biomarker as the independent variable. All the analyses were controlled for the confounding effects of age, sex and ASL sequence version. The distribution of NfL was skewed and required a logarithmic transformation. The results were considered statistically significant at p < 0.05. False discovery rate (FDR) correction was used to adjust for multiple comparisons.

### Voxel-wise analyses

In addition to the ROI-based analyses, whole-brain voxel-wise analyses were performed to compare CBF variations between groups and to assess the spatial associations of CBF with Aβ, tau and other CSF markers in both CU individuals and those on the AD spectrum. All voxel-wise analyses were controlled for age, sex and ASL sequence version. The CBF images were multiplied by a GM mask and subsequently smoothed with a Gaussian kernel of 4 mm FWHM. The analyses were performed using threshold-free, cluster-enhanced (TFCE) permutation statistics using FSL randomise command (https://fsl.fmrib.ox.ac.uk/fsl/fslwiki/Randomise) with 5000 permutations. Statistical significance was set at the family-wise error (FWE) corrected threshold of p < 0.05.

### Disease progression modeling

Subtype and Stage Inference (SuStaIn) was employed to characterize the progression of AD (***Young et al., 2018***) in terms of different biomarkers via the pySuStaIn package (https://github.com/ucl-pond/pySuStaIn). SuStaIn is an unsupervised machine learning approach that combines clustering and disease progression modeling to unravel the phenotypic and temporal heterogeneity of any progressive disease (***Young et al., 2018, 2020***). It simultaneously identifies subgroups of individuals with common phenotypes (subtypes) and distinct temporal progression patterns (stages). This data-driven approach requires inputs expressed as z-scores relative to a control population. It builds on event-based modeling (***Fonteijn et al., 2012; Young et al., 2014***) delineating the disease progression as a series of events each of which corresponds to a biomarker switching from a normal to an abnormal level. SuStaIn replaces the instantaneous normal to abnormal transition of events with a linear z-score model in which the trajectory of each subtype is described by linear accumulation of a biomarker from one z-score to another over different stages. Markov Chain Monte Carlo sampling is then used to quantify the uncertainty of the sequence ordering (***Young et al., 2018***) upon which a positional variance diagram can be visualized. Seven biomarkers that are most commonly associated with AD were fed into SuStaIn. They comprised CSF levels of Aβ42/40 and P-tau217, tau-PET deposition in early, temporal/limbic and late ROIs, a composite volumetric measure in AD signature regions i.e., bilateral entorhinal, inferior/middle temporal and fusiform cortices (***Ossenkoppele et al., 2018; Jack Jr et al., 2017***) and weighted mean CBF of regions having voxel-wise associations with tau-PET load in the early ROI (Figure 3 A) in the AD spectrum. Note that in the Aβ-negative CU individuals, the CBF values were extracted from the same voxel-wise correlation maps (see Figure 3 A). The values for each biomarker were represented as a z-score relative to Aβ-negative CU subjects who were younger than 50 years old (control population; N = 44), and thresholds were set at 1, 2 and 3 standard deviations. Except for being a reference for SuStaIn, the data from the control population were not included in any other analyses. Given that SuStaIn cannot handle missing data, participants with at least one missing value in any of the aforementioned markers were excluded. As such, the model was run on a sample of 203 individuals.

## Data Availability

Anonymized data will be shared by request from a qualified academic investigator for the sole purpose of replicating procedures and results presented in the article as long as data transfer is in agreement with EU legislation on the general data protection regulation and decisions by the Ethical Review Board of Sweden and Region Skåne, which should be regulated in a material transfer agreement.

## Acknowledgments

The authors thank the participants of the Swedish BioFINDER-2 study for their enthusiastic commitment.

## Funding

The present study was supported by the European Research Council (grant no. 311292), the Swedish Research Council (grants no. 2016-00906 and 2018-02052), the Knut and Alice Wallenberg Foundation (grant no. 2017-0383), the Marianne and Marcus Wallenberg Foundation (grant no. 2015.0125), the Swedish Alzheimer Foundation (grant no. AF-745911), the Swedish Brain Foundation (grant no. FO2019-0326), the Swedish federal government under the ALF agreement (grants no. 2018-Projekt0279 and 2018-Projekt0226) and the Strategic Research Area MultiPark (Multidisciplinary Research in Parkinson’s disease) at Lund University. F.B. was supported by the NIHR biomedical research Centre at UCLH.

## Competing interests

O.H. has acquired research support (for the institution) from AVID Radiopharmaceuticals, Biogen, Eli Lilly, Eisai, GE Healthcare, Pfizer, and Roche. In the past 2 years, he has received consultancy/speaker fees from AC Immune, Alzpath, Biogen, Cerveau and Roche. J.P. is an employee of Siemens Healthcare. All other authors report no competing interests.

## Supplementary materials

**Supplementary Table 1.** Significant between group differences in CBF adjusted for age, sex, ASL sequence version, and the GM volume.

| | Aβ-negative CU vs. Aβ-positive CI | Aβ-positive CU vs. Aβ-positive CI |
| --- | --- | --- |
| Lateral Parietal | $\beta = -0.043$<br>$p = 0.03$ | |
| Middle frontal | $\beta = -0.063$<br>$P_{FDR} = 0.025$ | |
| Superior lateral occipital | | $\beta = -0.063$<br>$p = 0.024$ |

**Supplementary Table 2.** Significant associations of CBF in the pre-defined ROIs with tau uptake in early, temporal and late ROIs in the AD continuum, adjusted for age, sex ASL sequence version, and CSF levels of Aβ42/40.

|  | Lateral parietal | Lateral temporal | Superior lateral occipital | Middle frontal |
| --- | --- | --- | --- | --- |
| Early ROI (SUVR) | $\beta = -0.046$<br>$P_{FDR} = 0.0314$ | $\beta = -0.053$<br>$P_{FDR} = 0.0265$ | $\beta = -0.083$<br>$P_{FDR} = 0.0059$ | |
| Temporal meta-ROI (SUVR) | $\beta = -0.057$<br>$P_{FDR} = 0.000231$ | $\beta = -0.063$<br>$P_{FDR} = 0.000141$ | $\beta = -0.089$<br>$P_{FDR} = 0.000044$ | |
| Late meta-ROI (SUVR) | $\beta = -0.119$<br>$P_{FDR} = 0.00012$ | $\beta = -0.089$<br>$P_{FDR} = 0.00451$ | $\beta = -0.137$<br>$P_{FDR} = 0.00069$ | $\beta = -0.084$<br>$P_{FDR} = 0.00736$ |

**Supplementary Table 3.** Relationship between CBF in the pre-defined ROIs and tau SUVR in early, temporal and late ROIs in the AD continuum, adjusted for age, sex, ASL sequence version, and cognitive status. Note that the observed associations between CBF and tau burden in the early ROI remained statistically significant after adjustment for the cognitive status but did not survive FDR correction.

|  | Lateral parietal | Lateral temporal | Superior lateral occipital | Middle frontal |
| --- | --- | --- | --- | --- |
| Early ROI (SUVR) | $\beta = -0.041$<br>$p = 0.0477$ | $\beta = -0.050$<br>$p = 0.024$ | $\beta = -0.065$<br>$p = 0.0161$ | |
| Temporal meta-ROI (SUVR) | $\beta = -0.055$<br>$P_{FDR} = 0.0013$ | $\beta = -0.065$<br>$P_{FDR} = 0.00056$ | $\beta = -0.080$<br>$P_{FDR} = 0.00056$ | |
| Late meta-ROI (SUVR) | $\beta = -0.114$<br>$P_{FDR} = 0.00083$ | $\beta = -0.081$<br>$P_{FDR} = 0.0179$ | $\beta = -0.108$<br>$P_{FDR} = 0.0179$ | $\beta = -0.071$<br>$P_{FDR} = 0.0392$ |

**Supplementary Table 4.** Available data per group and biomarker. Although the Aβ42/40 ratio values obtained by Lumipulse assay were used for stratification of the participants, they were excluded from the subsequent analysis on the association of CBF and CSF Aβ42/40. As such, the number of available data for this marker is less than the total number of the study participants.

| | Aβ-negative CU | Aβ-positive CU | Aβ-positive CI |
| --- | --- | --- | --- |
| Total N | 94 | 43 | 119 |
| ASL-CBF | 94 | 43 | 119 |
| Tau-PET | 94 | 43 | 119 |
| P-tau217 | 72 | 33 | 99 |
| Aβ-PET | 87 | 40 | 56 |
| Aβ42/40 ratio | 81 | 41 | 116 |
| NPTX2/T-tau ratio | 76 | 39 | 113 |
| NfL | 80 | 40 | 115 |

**Supplementary Figure 1.**
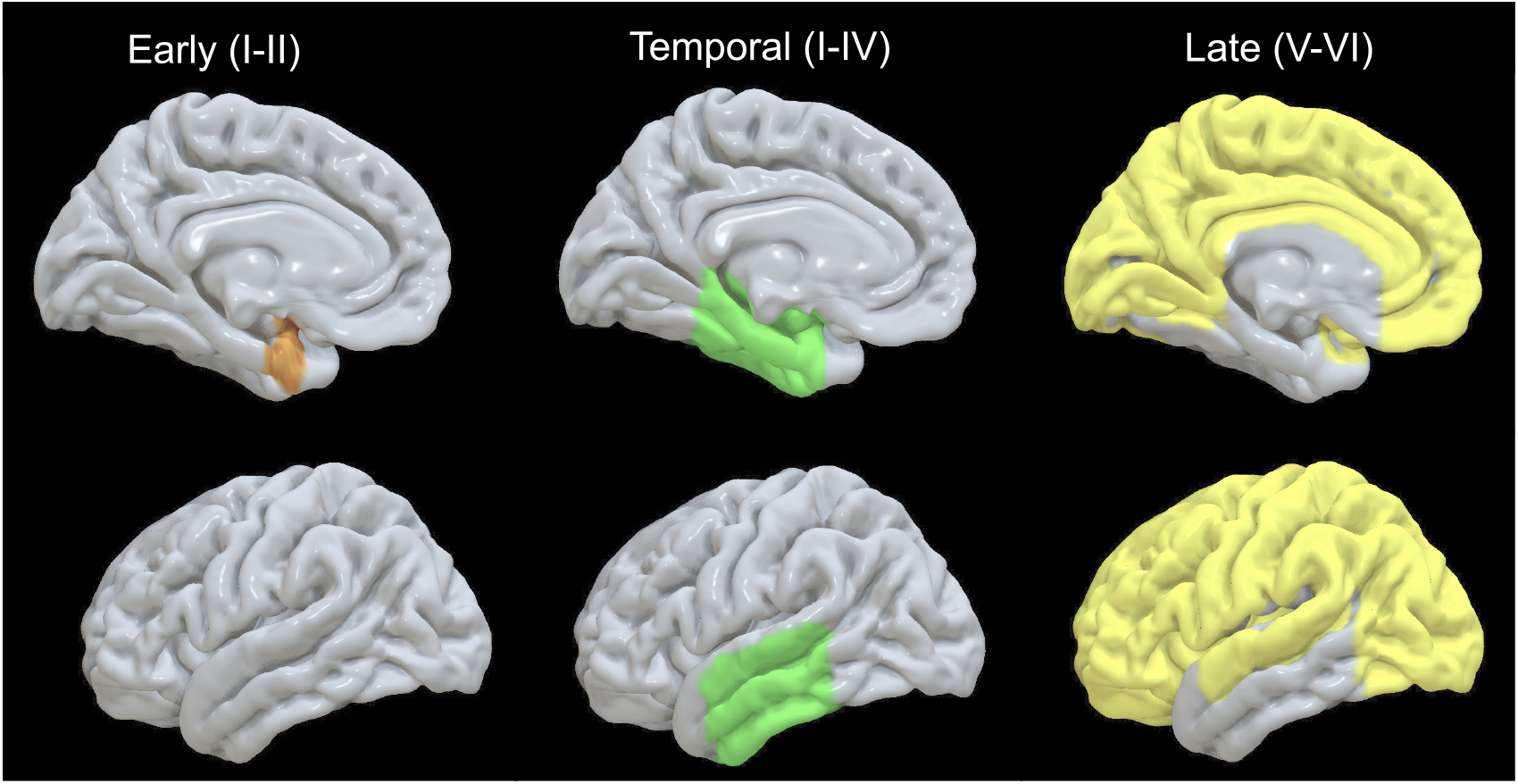
Tau-PET ROIs corresponding to Braak stages I-II, I-IV and V-VI (left, middle and right panels, respectively). Top and bottom panel display the lateral and medial view. For the sake of visualization, the bilateral ROIs are shown on the left hemisphere. The early ROI (I-II) consisted of entorhinal cortex while the remaining meta-ROIs encompassed the following sub-ROIs: I-IV (amygdala, fusiform gyrus, inferior/middle temporal gyri, parahippocampus and entorhinal cortex); V-VI (anterior/posterior cingulate, inferior/superior frontal, inferior/superior parietal, insular, lateral/medial occipital, lingual gyrus, middle frontal, orbitofrontal cortex, paracentral cortex, precentral/postcentral gyri, precuneus, superior temporal gyrus, and supramarginal gyrus).

**Supplementary Figure 2.**
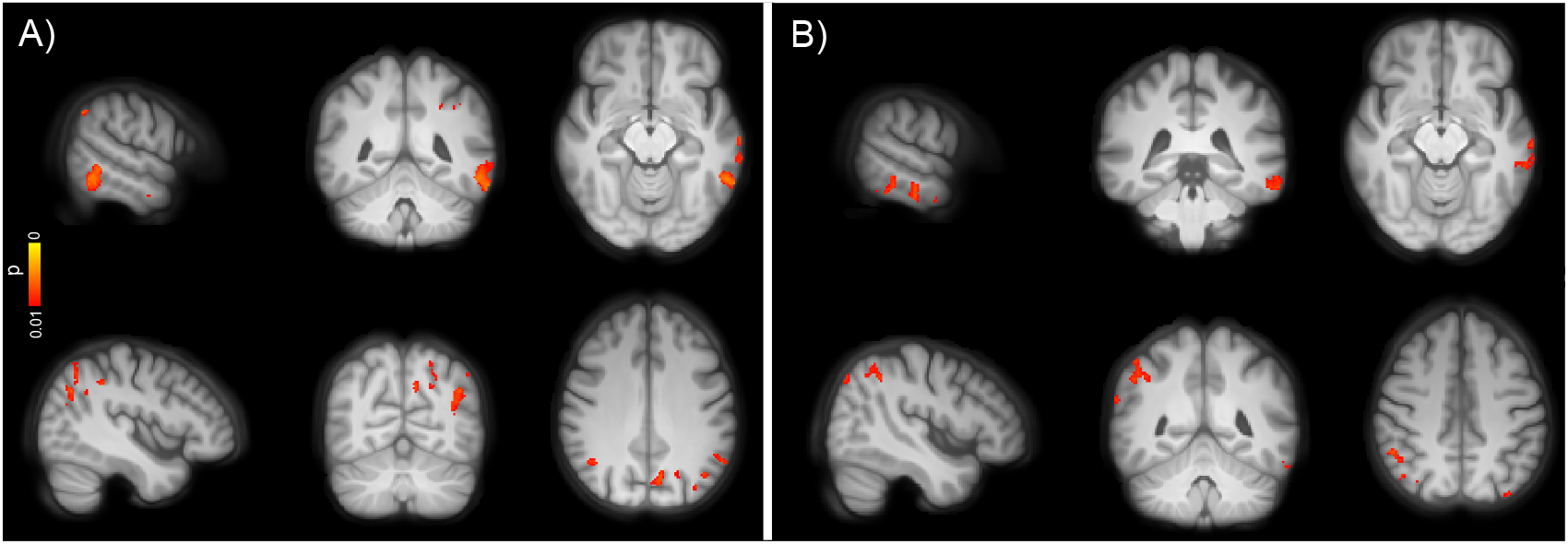
Voxel-wise group differences in CBF between A) Aβ-negative CU and B) Aβ-positive CU vs. Aβ-positive CI individuals corrected for age, sex, ASL sequence version and GM volume (p < 0.01, uncorrected). Conventions as for Figure 2.

**Supplementary Figure 3.**
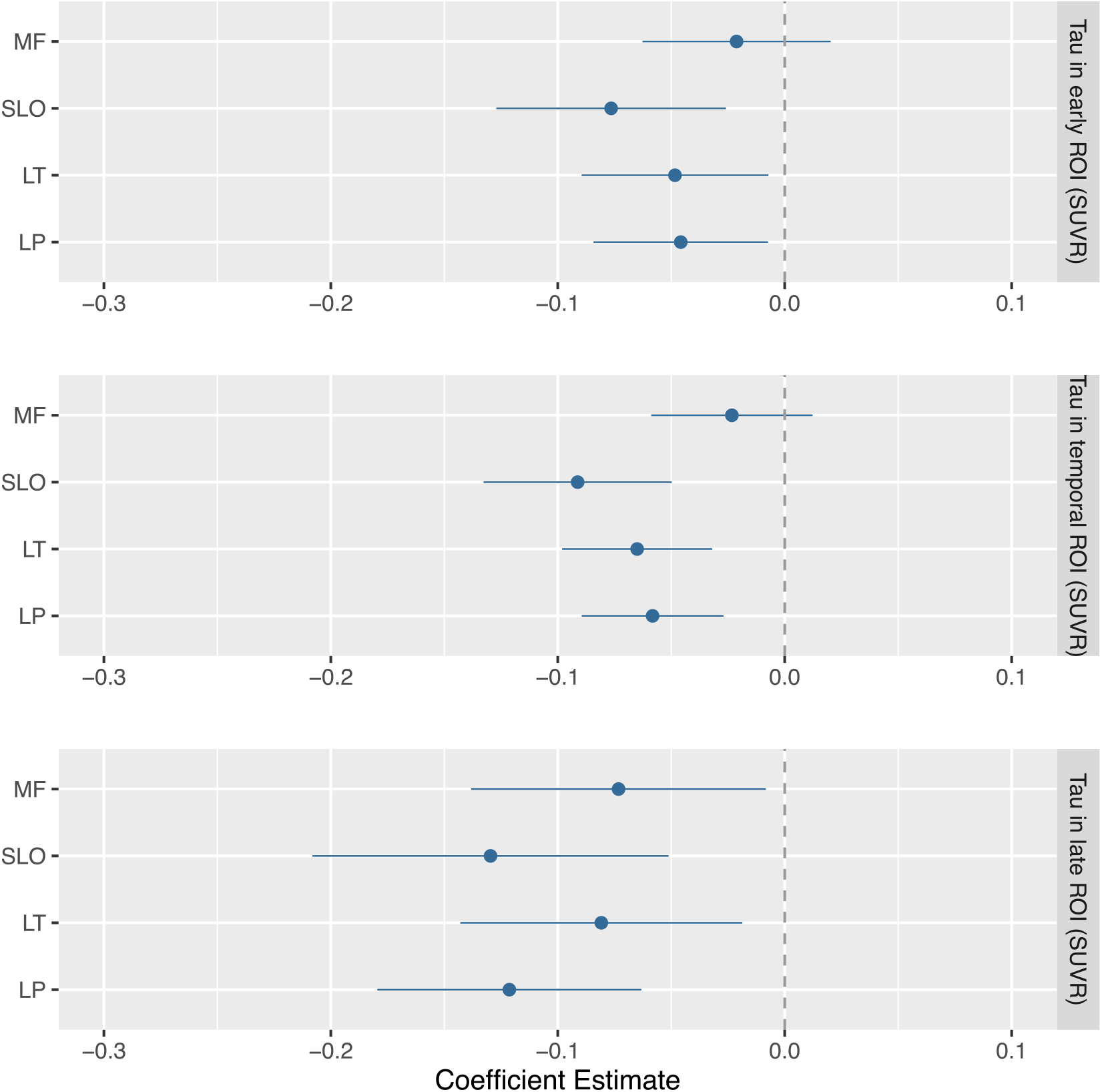
Regression coefficients (β) for the associations between CBF in a priori ROIs and the three tau-PET ROIs in the AD continuum, adjusted for age, sex, ASL sequence version, and GM volume. All the estimated coefficients are significant at *P*_*FDR*_ ≤ 0.05 except for the middle frontal being significant only in the late tau-PET ROI (bottom panel). LP = latera parietal, LT = lateral temporal, SLO = superior lateral occipital, and MF = middle frontal.

**Supplementary Figure 4.**
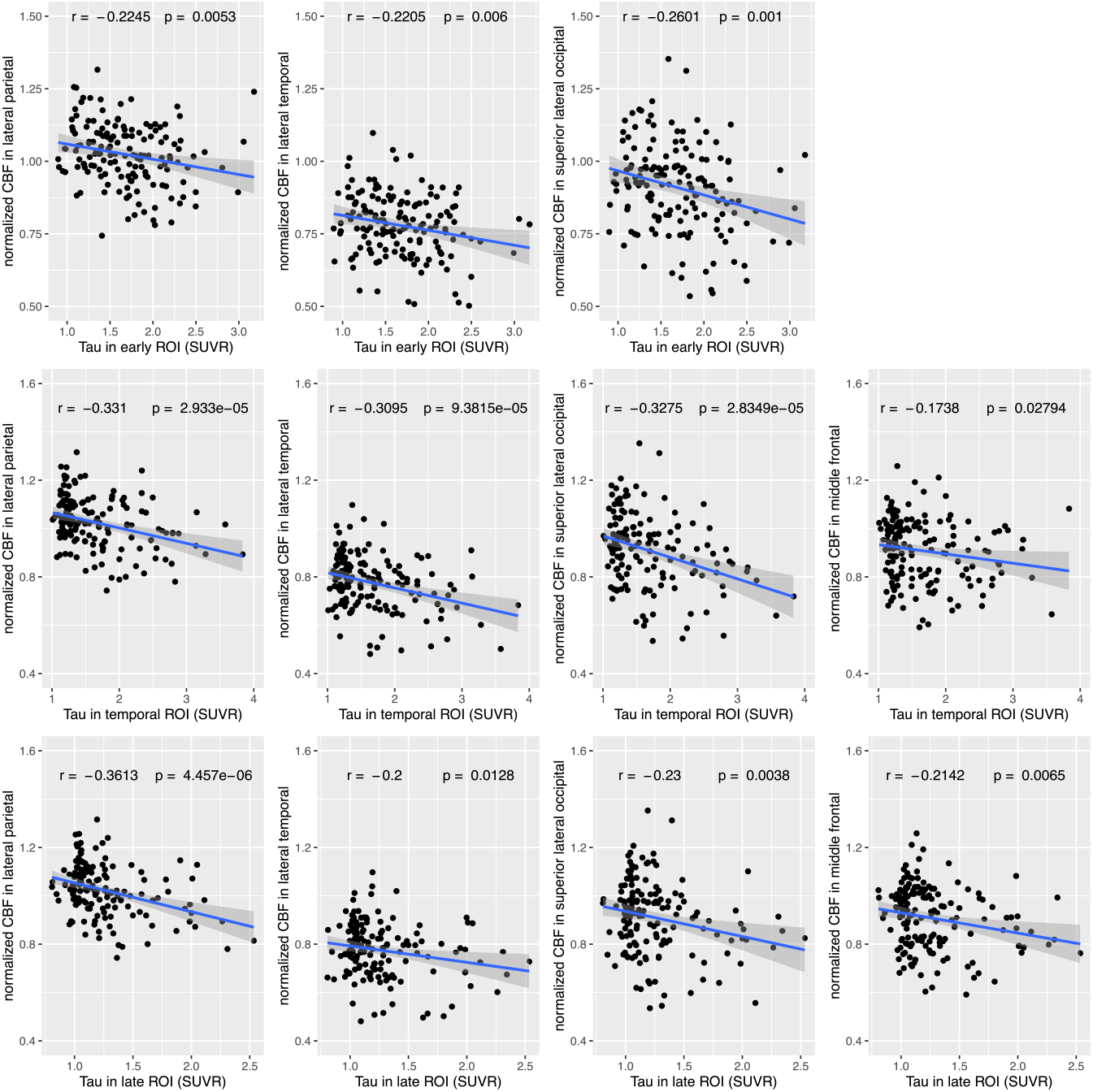
Overall significant associations between CBF in the pre-defined regions and tau load in early (top panel), temporal (middle panel) and late (bottom panel) ROIs in the AD continuum. The CBF in each pre-defined ROI is normalized to the CBF of the precentral gyrus (see Methods). The translucent area around the regression line indicates the 95% confidence interval for the regression coefficient. Note that the annotated statistical parameters (r and corresponding uncorrected p values) are based on the correlation of CBF and tau-PET load without adjusting for confounding variables.

**Supplementary Figure 5.**
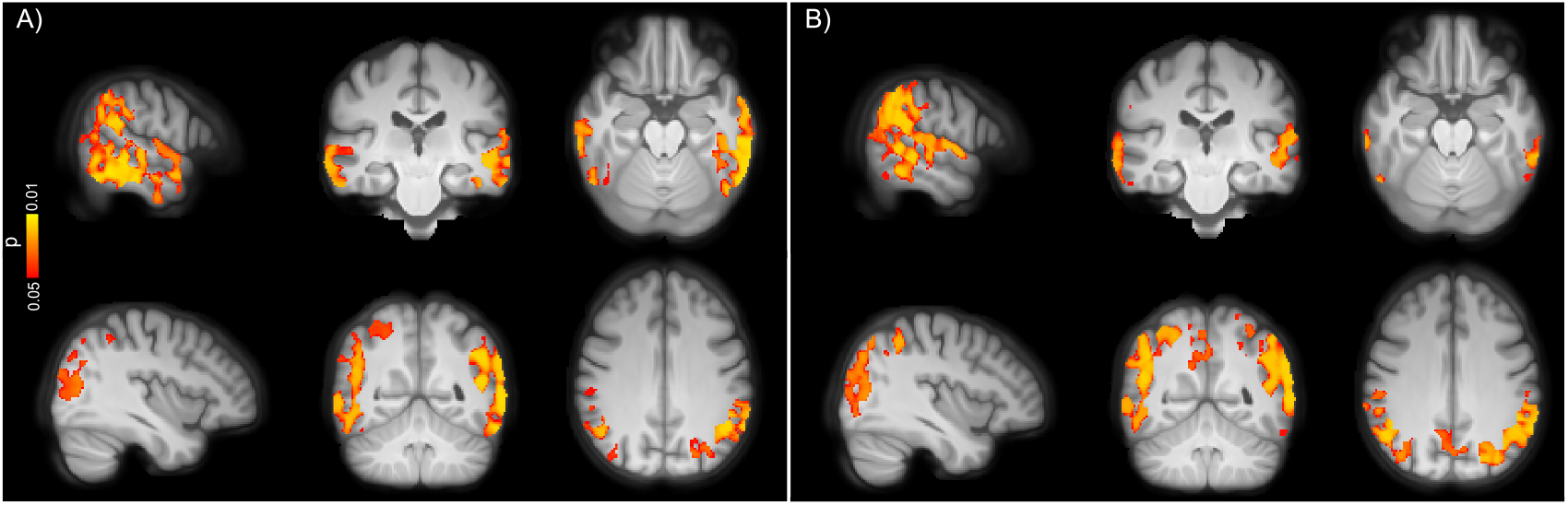
Voxel-wise associations between CBF and tau load in A) temporal and B) late meta-ROIs in the AD spectrum, corrected for age, sex, ASL sequence version and cognitive status (p < 0.05, FEW corrected). Clusters with markedly decreased CBF are visible in occipito-temporo-parietal ROIs. Conventions as for Figure 2.

**Supplementary Figure 6.**
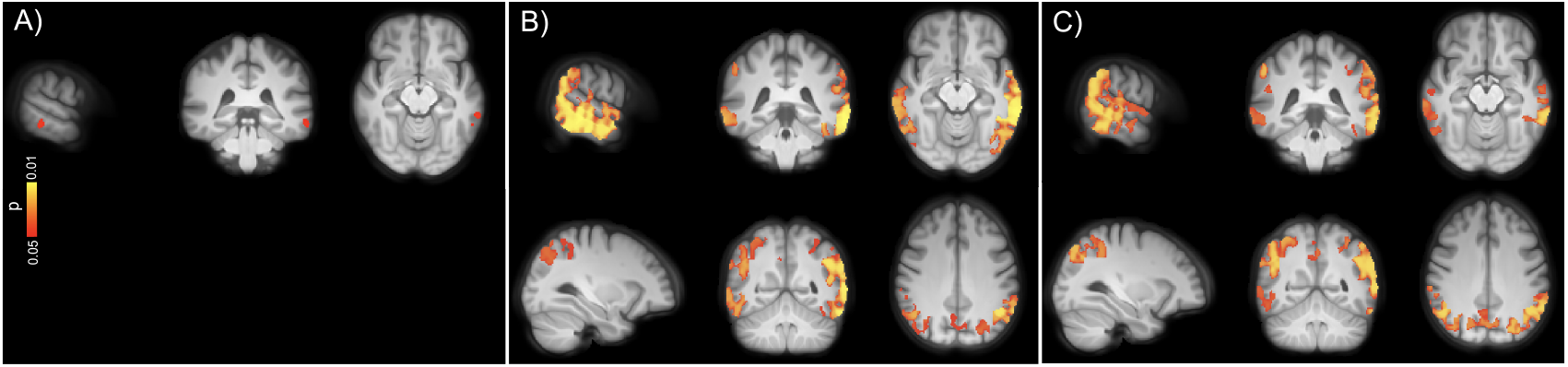
Voxel-wise associations between CBF and tau uptake in A) early B) temporal and C) late ROIs in the AD spectrum, adjusted for age, sex, ASL sequence version and GM volume (p < 0.05, FEW corrected). Conventions as for

**Supplementary Figure 7.**
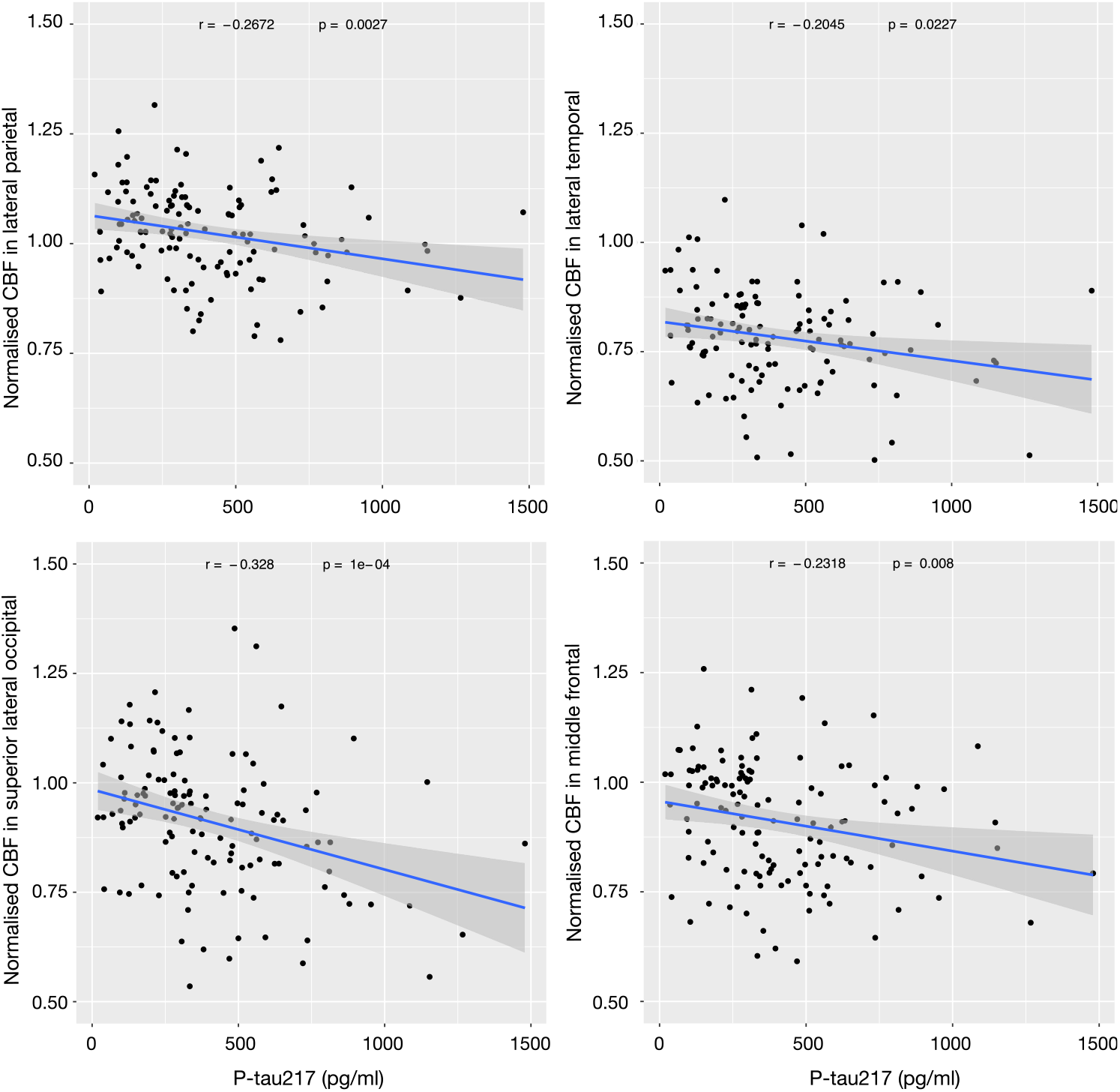
Higher P-tau217 is correlated with lower CBF in the AD spectrum. Conventions as for Supplementary Figure 4.

**Supplementary Figure 8.**
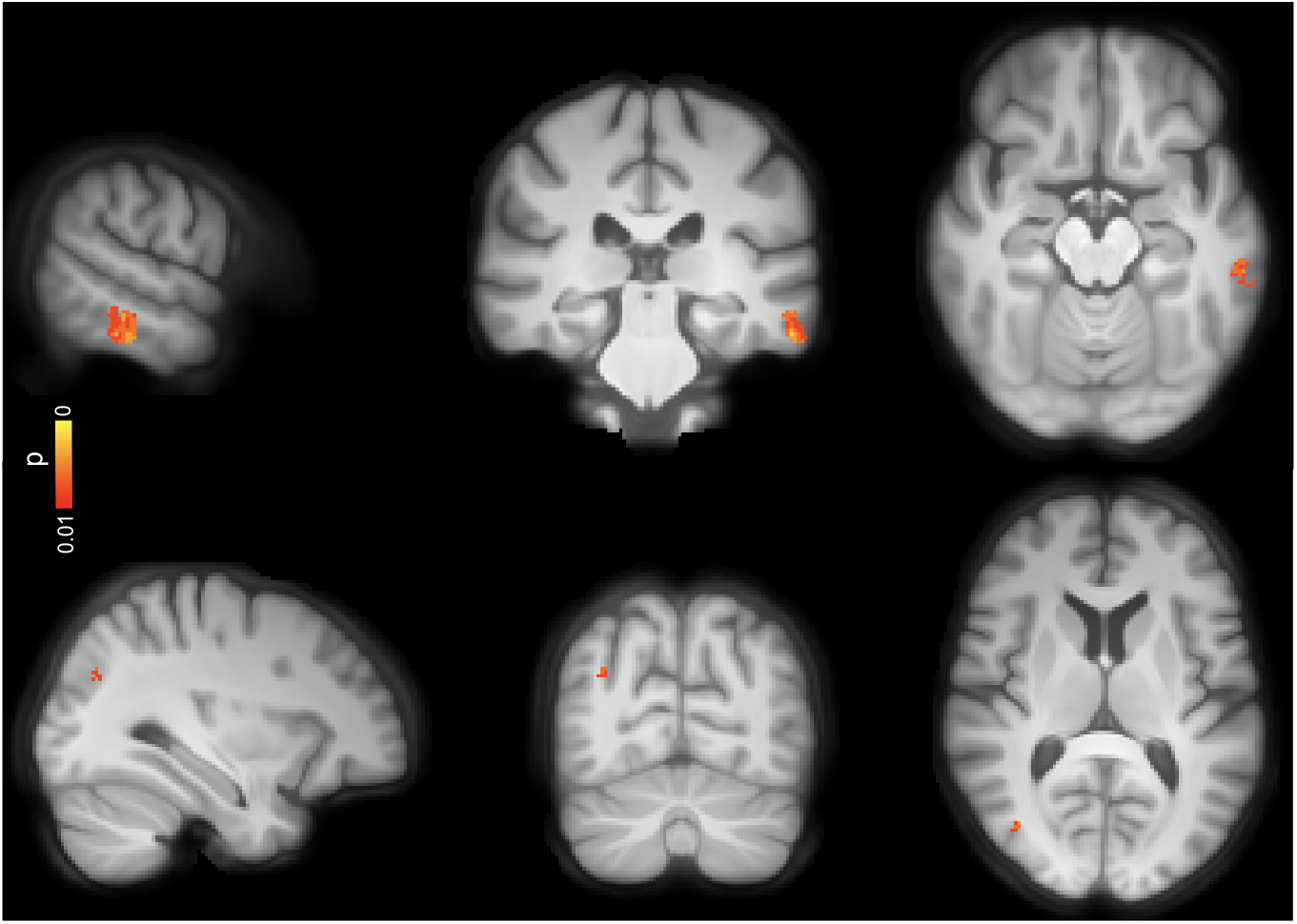
Voxel-wise associations between CBF and CSF levels of P-tau217 in the AD spectrum, corrected for ag, sex, ASL sequence version and GM volume. (p < 0.01, uncorrected). Conventions as for Figure 2.

**Supplementary Figure 9.**
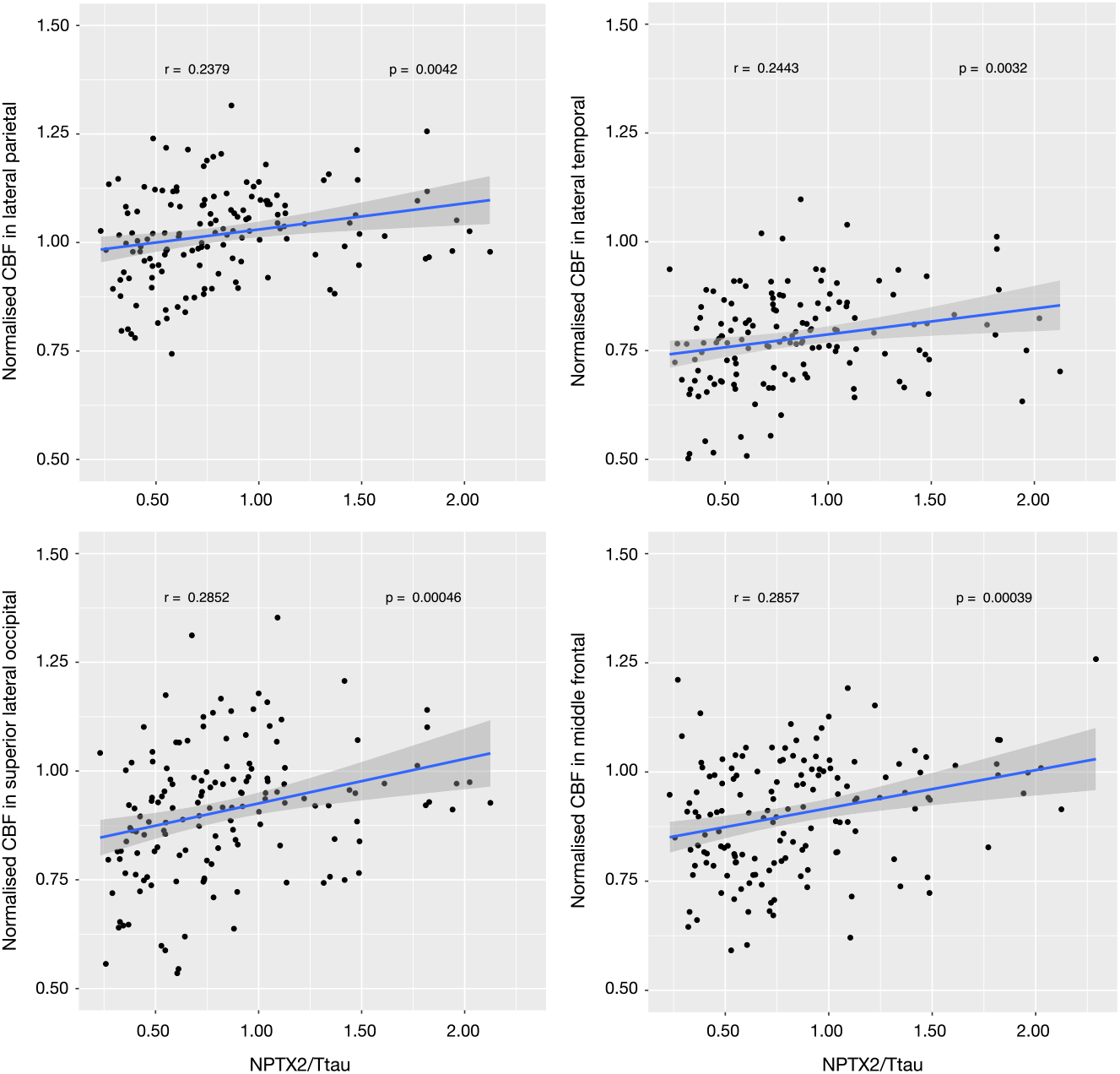
Lower NPTX2/T-tau is associated with lower CBF in the AD continuum. Conventions as for Supplementary Figure 4.

**Supplementary Figure 10.**
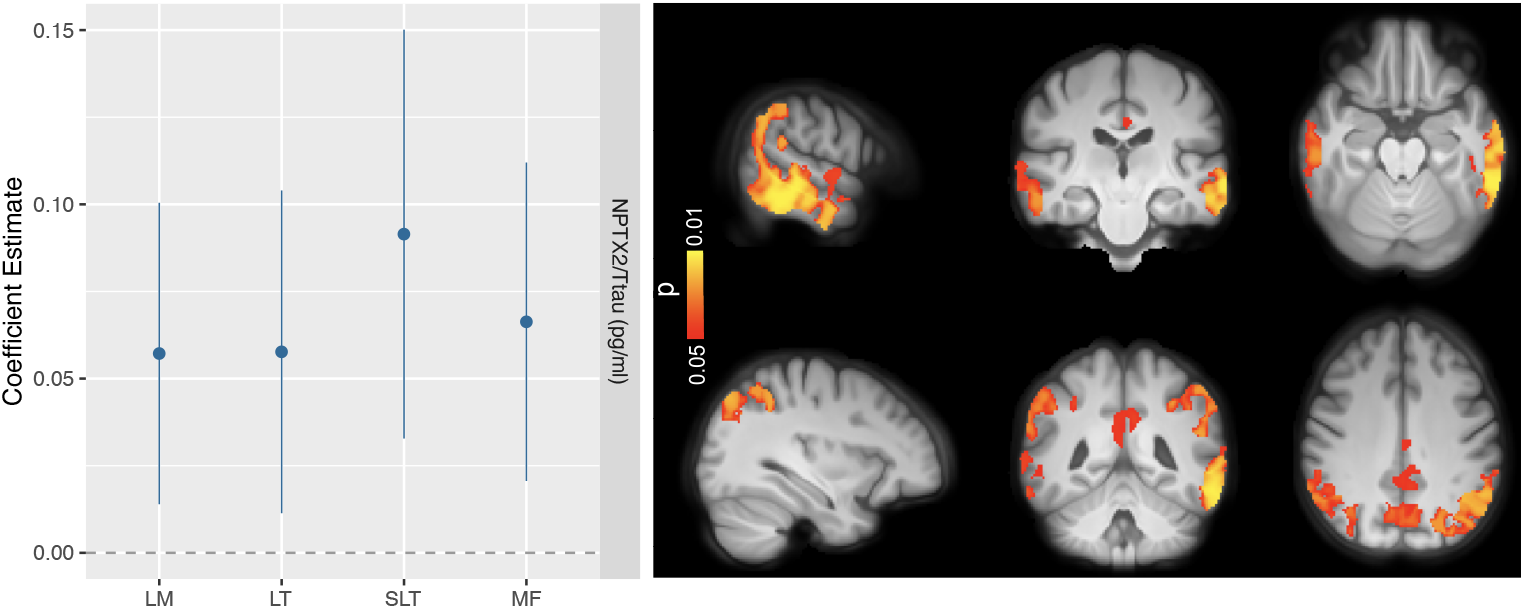
ROI-based and voxel-wise associations between CBF and NPX/T-tau in the AD continuum, corrected for age, sex, ASL sequence version and GM volume. The left panel presents the regression coefficients (β) all of which are significant at *P*_*FDR*_ ≤ 0.05, conventions as for Supplementary Figure 3. The voxel-wise associations (right panel) are significant at p < 0.05, FEW corrected.

## Notes

### Clinical Trial

NCT03174938

### Author Declarations

All participants gave written informed consent, and the study was approved by the Ethical Review Board in Lund, Sweden.

